# Incidence and outcome of intracerebral hemorrhage in a prospective cohort and healthcare data

**DOI:** 10.64898/2026.11.24.26368917

**Authors:** Alice Hosking, Neshika Samarasekera, James JM Loan, Tom J Moullaali, Mark A Rodrigues, Kristiina Rannikmäe, Colin Smith, Sohan Seth, William N Whiteley, Rustam Al-Shahi Salman, the Lothian Audit of the Treatment of Cerebral Haemorrhage Collaborators

**Author notes:** **Corresponding author:** Alice Hosking, Address: 49 Little France Crescent, Edinburgh EH16 4SB.

## Abstract

**Background:** The global burden of intracerebral hemorrhage (ICH) remains high. Identifying trends in incidence and outcome requires resource-intensive epidemiological surveillance. Our objective was to characterise incidence, survival and functional outcome of people with ICH in a cohort study; and compare to healthcare systems data (HSD) in the same region.

**Methods:** We conducted a prospective population-based inception cohort study in Lothian, Scotland, UK (the Lothian Audit of Treatment of ICH, LATCH). LATCH included all spontaneous non-traumatic ICH in Lothian residents ≥16 years between 1^st^ June 2010 – 31^st^ May 2013 and 1^st^ November 2018 – 31^st^ October 2021. We identified and characterized people with ICH in HSD in the same region and period using linked primary and secondary care records, community prescriptions and death records. We calculated incidence per 100,000 person-years; 30-day survival; and sensitivity and positive predictive value of ICH detection in HSD. For functional outcome one year after ICH, we compared the modified Rankin Scale score (mRS) in LATCH to days alive and out of hospital in HSD.

**Results:** In LATCH, from 1283 possible cases, 1021 were first-ever ICH. We identified 876 people with ICH in HSD in the same period. In LATCH, ICH incidence was 23.7 (95%CI 22.2–25.2) per 100,000 person-years (48.5% male, mean age 72 (SD 14.6) years), with no time trend (trend in crude incidence rates 0.31/year (−3.78–4.39)). ICH incidence in HSD was 23.1 (21.6–24.6) per 100,000 person-years (48.3% male, mean age 74.2 (SD 14.0) years). The positive predictive value of ICH identified in HSD versus LATCH was 78.3% (75.4%–81.0%). 30-day case fatality was 41.2% (38.1%–44.2%) in LATCH and 39.0% (35.5%–42.3%) in HSD. We found good correlation between functional outcome measures (Spearman’s r=0.88, p<0.001).

**Conclusions:** The burden of ICH remains persistently high. HSD appears accurate for assessing incidence, survival and functional outcome.

## Introduction

Intracerebral hemorrhage (ICH) accounts for a quarter of strokes worldwide,^1^ but continuous accurate measurement of disease incidence and outcome is resource-intensive.

Population-based cohort studies, largely conducted in higher income countries, provide estimates of ICH incidence at particular time-points,^2^ but changing population demographics and behaviours affect incidence. National stroke registries such as SSNAP (England), the Scottish Stroke Care Audit, Riks-stroke (Sweden) and the Ontario Stroke Register maintain continuous recording of cases, although are resource intensive as information is entered by hand.^3–5^

Healthcare systems data (HSD), routinely collected when people make contact with health services, provide an opportunity to ascertain and monitor cases for research and audit purposes; characterise underlying comorbidities in both those with disease and unaffected populations; and describe outcomes.^6^ However, valid conclusions depend on accurate case identification and characterisation.^7–10^ The optimal validation study for HSD would compare both primary and secondary care data to a reference standard of a prospective population-based cohort.

Therefore, we aimed to characterise the incidence, characteristics, survival and functional outcome of people with ICH identified in a population-based inception cohort study in the Lothian region of Scotland; and compare to those identified in a healthcare systems data resource, DataLoch, which includes primary care, secondary care, death and prescriptions data for the same region.

## Methods

### Setting

In Scotland, almost all healthcare is provided by the National Health Service (NHS). Lothian health board comprises the City of Edinburgh and surrounding areas, with three acute hospital sites. Hospital admissions data is coded from the contents of the discharge letter and entered into a central database (Scottish Morbidity Record (SMR) 01). Primary care physicians maintain a separate record of medical conditions including those diagnosed in hospital. We obtained population estimates from the Scottish Government and the UK Office for National Statistics.^11,12^

### Population-based inception cohort

The Lothian Audit of the Treatment of Cerebral Hemorrhage (LATCH) was a prospective, population-based inception cohort study of people with ICH confirmed by brain imaging or pathology, in the Lothian region (mid 2010 population ≥16 years 684,880), in two distinct three-year cohorts (1^st^ June 2010 – 31^st^ May 2013, and 1^st^ November 2018 – 31^st^ October 2021). The study design has been described previously.^13^

ICH cases were defined as symptomatic spontaneous (i.e. non-traumatic) ICH in people ≥16 years resident in Lothian. Cases were identified prospectively from multiple sources, including liaison with clinical teams, daily screening of all inpatient and outpatient computed tomography brain imaging, and screening of ward admissions. Retrospective methods identified cases missed from prospective pursuit: review of an automatic report of ICD-10 codes entered by nursing staff on wards, sudden deaths held by the Office of the Procurator Fiscal, the Scottish Stroke Care Audit and hospital admissions ((SMR01) held by the Public Health Scotland. Underlying pathology was ascertained from discussion with clinical teams and multidisciplinary neuroradiology meetings, radiology and post-mortem reports and clinical case notes. Macrovascular included arteriovenous malformations, cavernous malformations, cerebral venous sinus thrombosis, arterial aneurysm and dural arteriovenous fistula. Malignancy included primary and secondary solid tumours, and haematological malignancy. If no pathology was recorded, we presumed microvascular. Follow-up was by annual questionnaire sent to the primary care physician, and death records from National Records of Scotland.

In Lothian, hospital staff can view a summary of the primary care record. LATCH investigators combined this with inpatient notes to identify comorbidities and current medication. Hypertension was defined as either documented history of hypertension or taking antihypertensive medication at time of ICH.

In LATCH, ICH incidence was calculated by dividing the number of ICH cases in each full year of LATCH by the mid-year population estimate for all Lothian residents aged ≥16 years, for the calendar year with the greatest overlap. We then reweighted the estimates for the whole 6-year period to the age and sex distribution of UK population ≥16 years in 2022. We calculated a time trend using linear regression.

### Healthcare systems data

DataLoch is an HSD resource that provides access to pseudonymised person-level routine hospital, primary care, prescription and death records in Lothian; 88% of Lothian residents are registered at a participating practice. Within HSD, we obtained hospital, prescription and primary care records for all people aged ≥16 years from 1^st^ January 2000 to 31^st^ December 2022.^14^ For the periods of LATCH, we identified people with ICH in records of hospital admissions (SMR01) and death records using ICD-10 codes; and in primary care records using Read codes (code lists in Supplement 2). LATCH was linked to HSD using the Scottish Community Health Index number, which is unique for each person in Scotland. For the comparison with HSD, we only included people in LATCH with a record in HSD.

### Accuracy of HSD for ICH diagnosis

In hospital admission and death records, multiple ICD-10 codes can be assigned to each admission/death, with one identified as main condition/cause of death; in primary care records there is no hierarchy of codes. We identified ICH in HSD through either 1) primary care ICH phenotype (defined by DataLoch using Read codes, Supplement 2); 2) I61 ICD-10 code, anywhere in SMR01 or death records; 3) I61 ICD-10 code as main condition/cause of death in SMR01 or death records; and combinations of these. We defined LATCH participants with an HSD record as the gold standard. True positives were present in both LATCH and HSD; false positives were present only in HSD; and false negatives were present only in LATCH. We calculated the sensitivity (true positives/total in LATCH) and positive predictive value (true positives/(true positives + false positives)) of each method of ascertaining ICH diagnosis in HSD. There are no ICD-10 codes for pathological subtypes of ICH, so we identified cases of concurrent brain malignancy using ICD-10 codes within 1 year of index ICH, and intracranial aneurysms using procedure codes and ICD-10 codes within 6 months of index ICH. We identified head trauma codes on the same admission as ICH (Supplement 2).

### Accuracy of HSD for ICH incidence and characteristics

In HSD, we calculated ICH incidence for each year of the LATCH study and multiplied population estimates by 0.88 to account for coverage of the Lothian population.

We identified comorbidities with previously curated code lists in hospital, primary care and prescription data.^14,15^ Hypertension was defined as record of hypertension phenotype or prescription of antihypertensive medicine in the two months prior to ICH (Supplement 2).

To maximize capture of chronic kidney disease, in HSD we accessed eGFR from routine blood samples. We used a previously validated pragmatic electronic phenotype for CKD, with the most recent eGFR of <60ml/min/1.73m^2^, with at least one other value <60ml/min/1.73m^2^ more than 90 days prior.^16^ We recorded recent (in the two months prior to ICH) prescription of statin, anticoagulant and antiplatelet medicines.

We compared continuous variables between the LATCH cohort and people with ICH in HSD with two-sided student’s t-tests and discrete variables with chi-square tests.

### Accuracy of HSD for outcome ascertainment

We constructed Kaplan-Meier plots for time to death in the LATCH cohort compared to those with ICH in HSD. We compared survival between the two cohorts with Cox proportional hazards model, with the source dataset as a covariate, censored at end of follow-up time (31^st^ December 2022). We calculated days alive and out of hospital (DAOH) for each person in both HSD and LATCH. For each category of the modified Rankin Scale (mRS) score at 1 year obtained from annual primary care questionnaire follow-up in LATCH, we calculated the median DAOH.

### Regulatory approval

LATCH was approved by the NHS Lothian Caldicott Guardian. Patients in NHS Lothian were informed about use of their data for audit, and information leaflets were distributed to inform patients and their carers about their right to opt out.

The HSD project received approval through DataLoch, including: Caldicott Guardian approval, The Academic and Clinical Central Office for Research and Development sponsorship (AC24054), favourable ethical opinion under DataLoch’s delegated authority (reference 22/NS/0093), and public value assessment by the DataLoch Public Reference Group.

## Results

### Population-based inception cohort

LATCH identified 1072 people with ICH, of whom 1021 had a first-ever event in the time period, of whom 869/1021 (85.1%) had ICH presumed due to microvascular pathology (e and Table 1). Most of the remainder had either macrovascular pathology (95/1021; 9.3%) or malignancy (53/1021; 5.2%). The incidence of ICH in LATCH was 23.7 (95%CI 22.2 – 25.2) per 100,000 per year (48.5% male, mean age 72 (SD 14.6) years). ICH incidence increased dramatically with age, from 3.2 (2.7–3.6) per 100,00 per year in those <55 years to 123.2 (95%CI 106.6 – 141.8) per 100,000 in those 85 years and over. There was no significant difference in incidence between men and women at any age (Figure 1).

**Figure 1:**
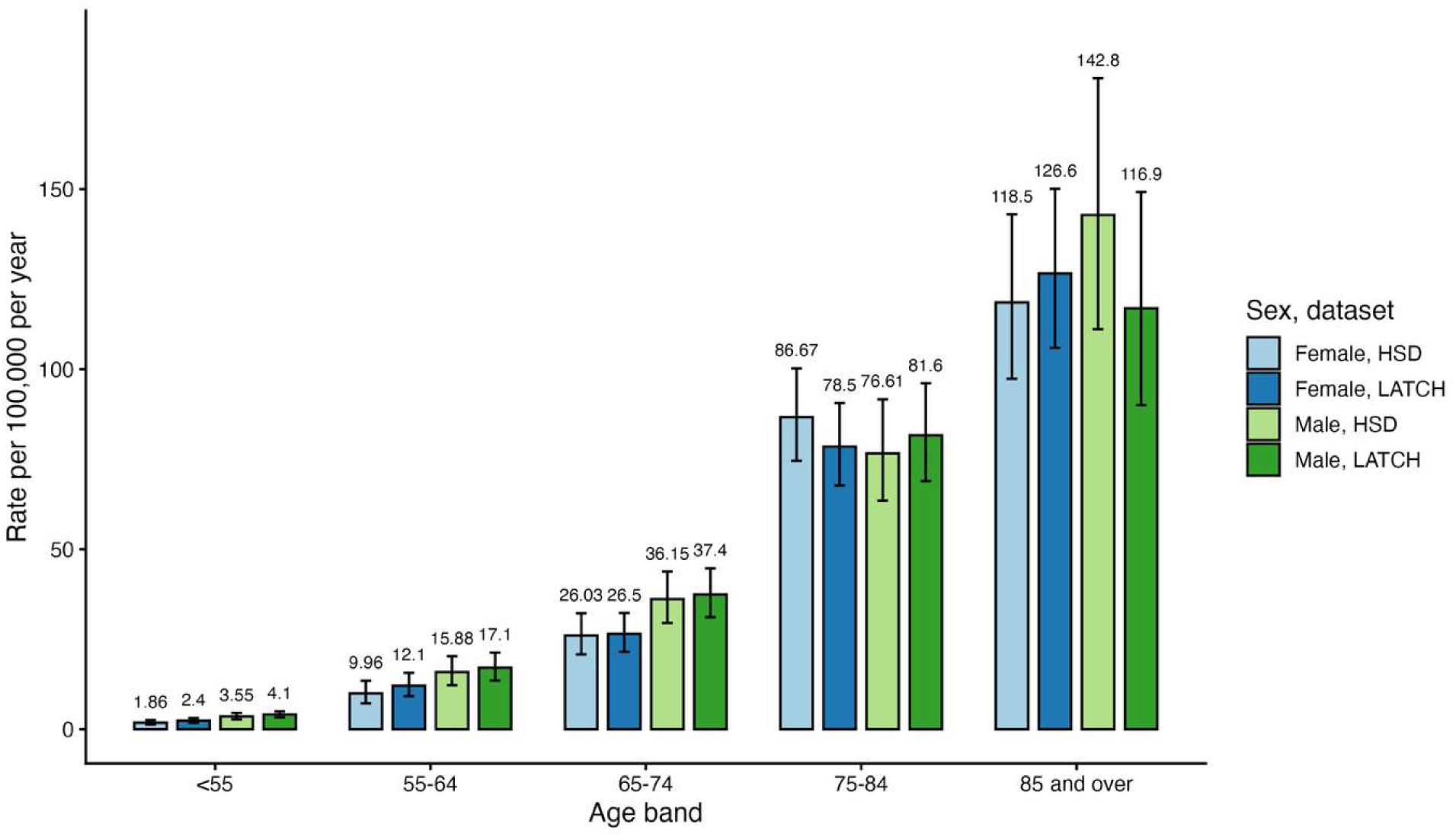
First-ever intracerebral hemorrhage (ICH) incidence, stratified by age, sex, and data source. LATCH = Lothian Audit of Treatment of Intracerebral Hemorrhage, a prospective, population-based inception cohort study of adults ≥16 years with ICH; HSD = healthcare systems data, comprising all adults in the Lothian region registered with a participating primary care physician.

**Table 1:** Underlying pathology of first-ever intracerebral hemorrhage (ICH) in the prospective, population-based incidence study (LATCH).

|  | **2010 – 2013 cohort**<br>(n=494) | **2018 – 2021 cohort**<br>(n=527) | **Overall**<br>(n=1,021) |
|---|---|---|---|
| Presumed microvascular | 417 (84.4%) | 452 (85.8%) | 869 (85.1%) |
| Macrovascular | 47 (9.5%) | 48 (9.1%) | 95 (9.3%) |
| Malignancy | 29 (5.9%) | 24 (4.6%) | 53 (5.2%) |
| Infective | 1 (0.2%) | 2 (0.4%) | 3 (0.3%) |
| Coagulopathy | 0 (0%) | 1 (0.2%) | 1 (0.1%) |

Macrovascular included arteriovenous malformations, cavernous malformations, cerebral venous sinus thrombosis, arterial aneurysm and dural arteriovenous fistula. Malignancy included primary and secondary solid tumours. Infective included abscess and encephalitis. Coagulopathy did not include anticoagulant administration. If no other pathology was identified, we presumed microvascular pathology.

We did not detect a trend in ICH incidence in Lothian across the time of the study (eFigure 3, time trend in crude incidence rates 0.31/year (95%CI −3.78 – 4.39)). Adjusted to the age- and sex-distribution of the UK population in 2024, the rate of first-ever spontaneous ICH of any cause was 28.1 (95%CI 27.6 – 28.5) per 100,000 person years; and the rate of first-ever ICH due to presumed underlying microvascular pathology was 24.4 (24.0 – 24.8) per 100,000 per year.

### Accuracy of HSD for ICH diagnosis

916/1021 people in LATCH with first-ever ICH had a primary care record in HSD, because DataLoch only includes 88% of primary care registrations in NHS Lothian. Of the 916 people in LATCH in HSD with first-ever ICH, 777 had presumed microvascular pathology.

Within the LATCH time periods (1^st^ June 2010-31^st^ May 2013 and 1^st^ November 2018-31^st^ October 2021) we identified 552 people in HSD with first-ever record of ICH in GP records; 876 in hospital and death records with I61 as main condition/cause of death; 1045 in hospital and death records with I61 in any position; and 1113 with I61 in hospital or death records or an ICH phenotype in GP records (Figure 2). The highest PPV of 78.3% (95%CI 75.4% – 81.0%) was obtained with I61 as main condition/cause of death in hospital and death records, without reference to primary care records, with sensitivity 74.9% (72.0% – 77.7%) (Table 2).

**Figure 2.**
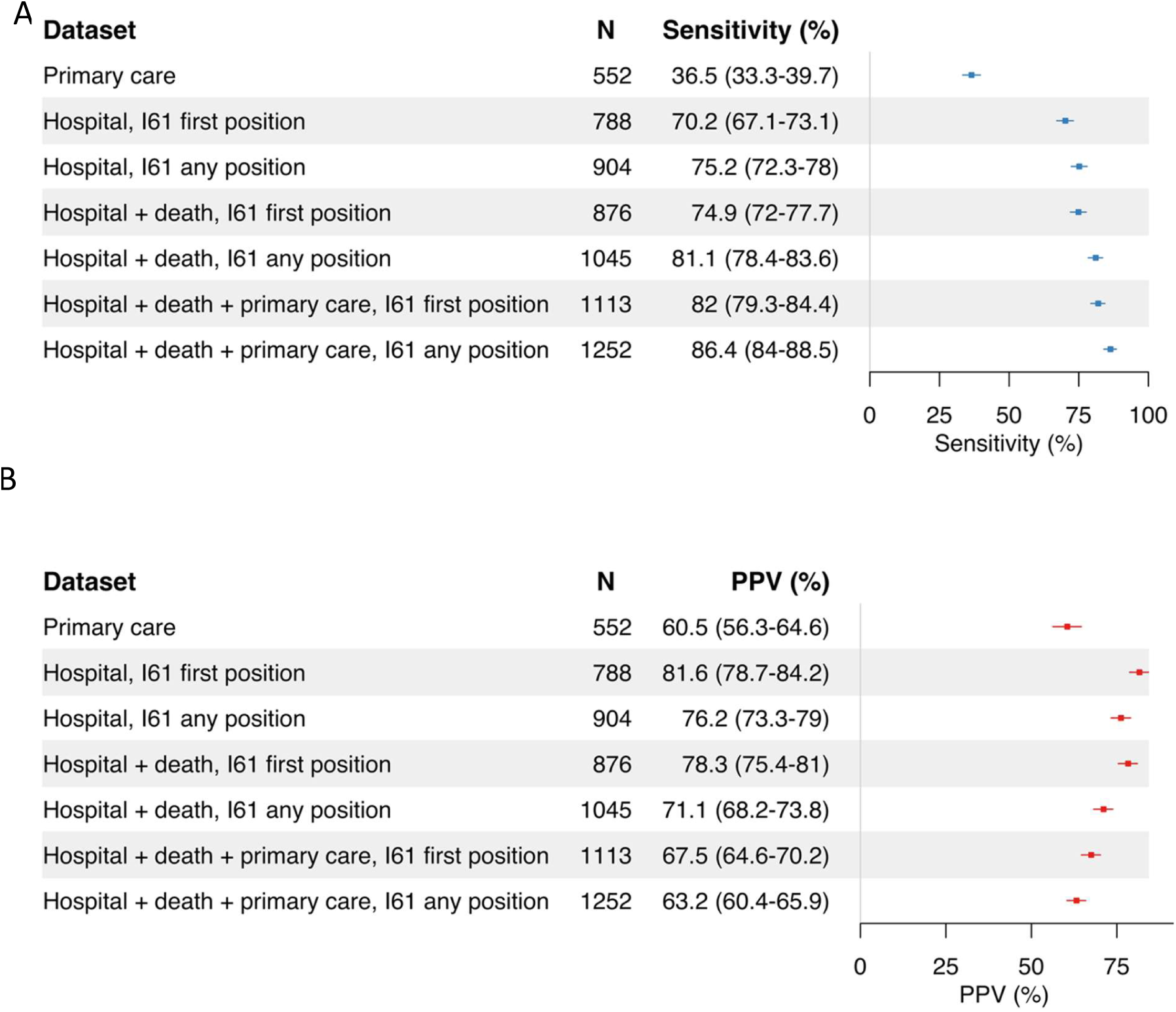
Positive predictive values (PPV) and sensitivity for identifying first-ever intracerebral hemorrhage (ICH) with healthcare systems data, using the prospective, population-based incidence study as a reference standard. Forest plots showing the sensitivity (A) and positive predictive value (PPV, B) of datasets and their combinations for identifying ICH in hospital admissions data (Scottish Morbidity Record 01) and death data using the ICD-10 code I61, and primary care data using a predefined list of Read codes (Supplement 2).

**Table 2:** Baseline characteristics of people with first-ever intracerebral hemorrhage (ICH) in LATCH and healthcare systems data (HSD) in the same period. We include the 916/1021 people in LATCH with a primary care record in HSD, because DataLoch only includes 88% of primary care registrations in NHS Lothian.

|  | **LATCH<br>(N=916)** | **HSD<br>(N=876)** | **p value** |
|---|---|---|---|
| **Demographics** |  |  |  |
| Age, mean (SD) | 71.7 (14.8) | 74.2 (14.0) | <0.001 |
| Sex, male (%) | 441 (48.1%) | 423 (48.3%) | 0.989 |
| **Medical conditions before ICH** |  |  |  |
| Hypertension | 509 (55.6%) | 565 (64.5%) | <0.001 |
| Diabetes mellitus | 114 (12.4%) | 128 (14.6%) | 0.203 |
| Atrial fibrillation | 176 (19.2%) | 187 (21.3%) | 0.287 |
| Dementia | 105 (11.5%) | 111 (12.7%) | 0.476 |
| Heart failure | 44 (4.8%) | 82 (9.4%) | <0.001 |
| Ischemic stroke or transient ischemic attack | 175 (19.1%) | 151 (17.2%) | 0.336 |
| Myocardial infarction | 65 (7.1%) | 89 (10.2%) | 0.0258 |
| Chronic kidney disease | 121 (13.2%) | 139 (15.9%) | 0.1103 |
| Pulmonary embolism | 35 (3.8%) | 34 (3.9%) | 0.947 |
| Deep vein thrombosis | 38 (4.1%) | 34 (3.9%) | 0.867 |
| Peripheral vascular disease | 36 (3.9%) | 52 (5.9%) | 0.0636 |
| Angina | 96 (10.5%) | 127 (14.5%) | 0.0123 |
| Epilepsy | 35 (3.8%) | 29 (3.3%) | 0.649 |
| Migraine | 65 (7.1%) | 27 (3.1%) | <0.001 |
| **Medicines prescribed <2 months before ICH** |  |  |  |
| Statin | 315 (34.4%) | 290 (33.1%) | 0.176 |
| Antiplatelet agent | 274 (29.9%) | 247 (28.2%) | 0.174 |
| Oral anticoagulant | 153 (16.7%) | 121 (13.8%) | 0.017 |
| **Smoking** |  |  |  |
| Current | 179 (19.5%) | 143 (16.3%) | <0.001 |
| Ex | 269 (29.4%) | 394 (45.0%) |  |
| Never | 392 (42.8%) | 294 (33.6%) |  |
| Missing | 76 (8.3%) | 45 (5.1%) |  |

The sensitivity of HSD for non-microvascular ICH using I61 as main condition/cause of death was 33.8% (95%CI 26.0% – 42.3%). HSD identified 32 cases with proximate malignancy/macrovascular codes; 23 of these were in LATCH; 20 had a malignancy/macrovascular cause in LATCH. Excluding people with malignancy/macrovascular codes from HSD and comparing only to those with a presumed underlying microvascular pathology gave a sensitivity 81.2% (78.3% – 83.9%) but reduced PPV to 73.7% (70.6% – 76.6%)).

The 230 false negatives (people with ICH in LATCH but not HSD) were younger than overall cohort (age 66.5 (SD 16.7) years, 45.9% male). The most common ICD-10 code was I62.9, “Intracranial hemorrhage (nontraumatic), unspecified”, the main condition/cause of death in 35 people’s death or SMR01 records. Fourteen people with ICH in HSD had head trauma codes on the same admission; <10 were false positives.

For all subsequent analyses we therefore defined ICH in HSD using I61 as main condition/cause of death in hospital and death records.

### Accuracy of HSD for ICH incidence and characteristics

Incidence of ICH in HSD was 23.1 (21.6–24.6) per 100,000 per year and similar to the LATCH cohort at all ages (Figure 1). People in LATCH were slightly younger than those in HSD with ICH (mean LATCH age 71.7 years (SD 14.8), vs 74.2 (14.0) years), Table 2). The proportion of men was similar (48.1% vs 48.3%). Compared to LATCH, using HSD to define people with ICH and their medical history identified higher proportions with: heart failure (LATCH, 4.8% vs HSD, 9.4%), peripheral vascular disease (3.9% vs 5.9%), hypertension (55.6% vs 64.5%), previous MI (7.1% vs 10.3%), chronic kidney disease (13.2% vs 15.9%) and angina (10.5% vs 14.5%); and a lower proportion with migraine (7.1% vs 3.1%)

### Accuracy of HSD for outcome ascertainment

In LATCH, 30-day case fatality from date of symptom onset was 41.2% (95%CI 38.1% – 44.2%) for all pathological subtypes; 42.2% (38.9% – 45.4%) for those with presumed microvascular pathology; and 35.5% (27.5% – 42.7%) for those with identified underlying pathology (eFigure 4 and eTable 1 in Supplement 1). Results were very similar if the start date was the date of initial CT scan. For the 876 people with ICH in HSD, 30-day case fatality from date of admission was 45.1% (41.7% – 48.3%), or 39.0% (35.5%-42.3%) if only hospital admission were used to identify cases. Kaplan Meier curves show shorter survival for HSD compared to the LATCH cohort (Figure 3).

**Figure 3:**
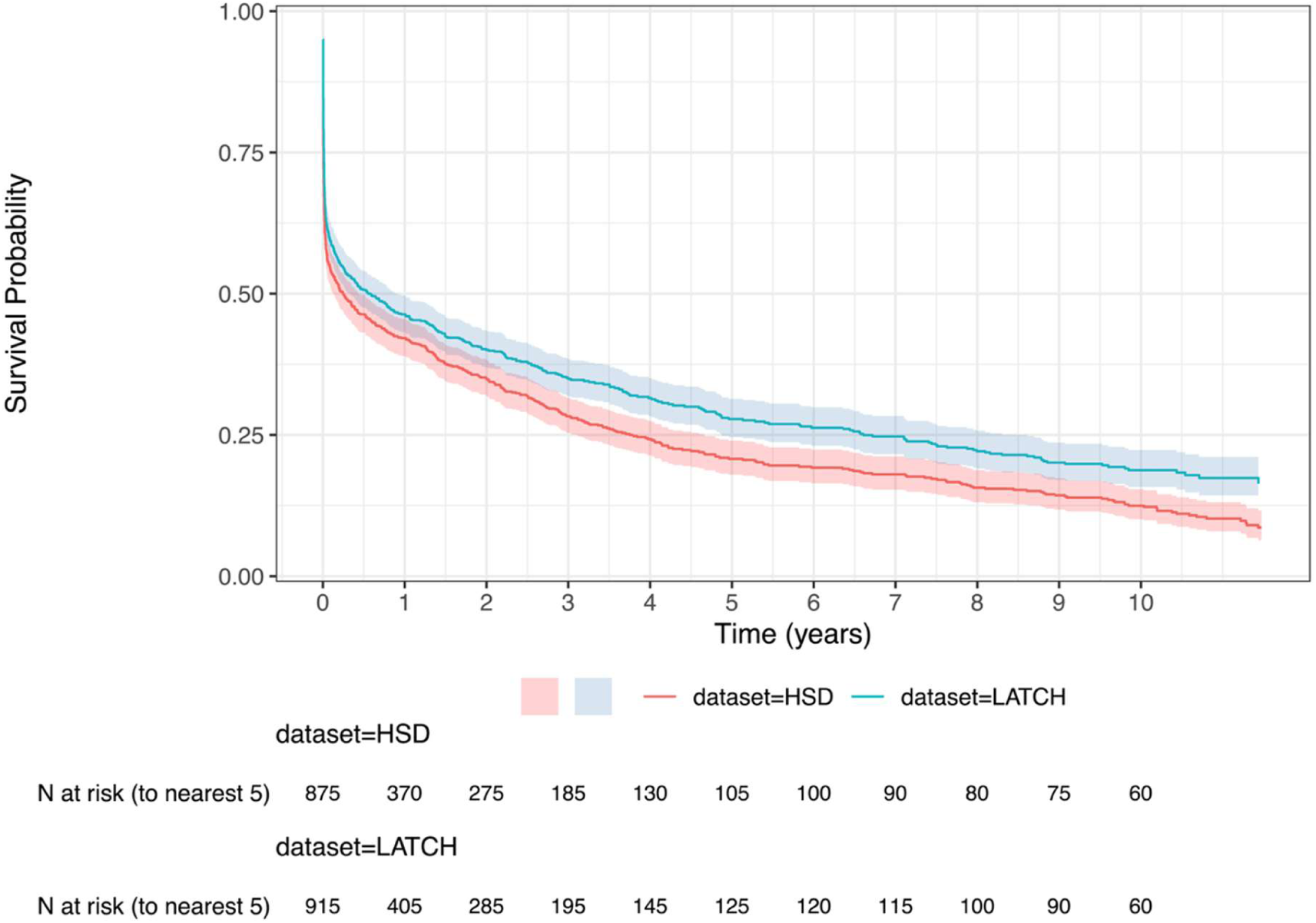
Kaplan-Meier survival curves of unadjusted time to death for the LATCH cohort compared to everyone with first-ever intracerebral hemorrhage (ICH) in HSD. ICH in HSD was identified using hospital admission and death data; we compare to the 916 first-ever ICH in LATCH with a primary care record in HSD. Numbers are given to the nearest 5 for data suppression.

The mortality HR for LATCH compared to HSD was 0.81 (95%CI 0.73 – 0.90), indicating shorter time to death for ICH in HSD. When LATCH was compared to ICH in HSD identified only in hospital admissions, the HR was non-significant at 0.91 (0.81 – 1.09), suggesting the main source of difference in survival was people identified only at death (Figure 3, panel B). Another source of difference was people identified only on second or subsequent admission after ICH, who therefore had shorter apparent survival times in HSD.

### Comparing functional outcome

In LATCH, one year after ICH, 226/1021 (22.1% (95%CI 19.3%–25.2%) had mRS 0 – 2 (independent); 755/1021 (73.9% (68.8%–79.4%) had mRS 3 – 6 (dead or dependent); 40 were not recorded. For those with presumed underlying microvascular pathology, 181/869 (20.1%) had mRS 0 – 2; 655/869 (78.6% (69.7%–81.4%)) had mRS 3 – 6; 33 were not recorded. For all first-ever ICH in the LATCH cohort, correlation between 1-year mRS in LATCH and DAOH calculated from HSD was 0.60 (p<0.001), excluding mRS 6 (dead). Including mRS 6 at one year, correlation was 0.88 (p<0.001) (Figure 4).

**Figure 4:**
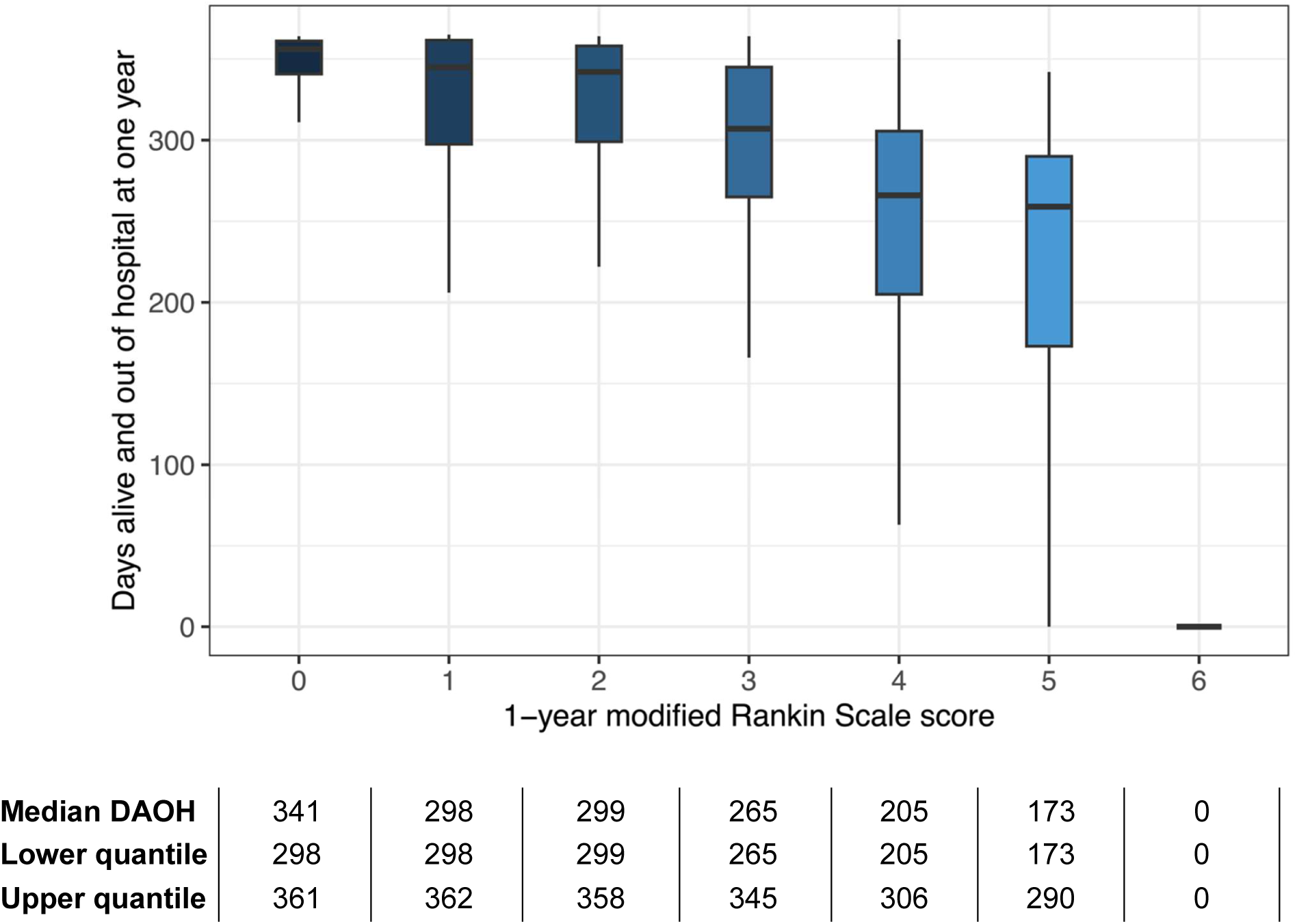
Box-plot comparing days alive and out of hospital in HSD to modified Rankin Scale score in LATCH, one year after first-ever intracerebral hemorrhage (ICH). Correlation between days alive out of hospital and modified Rankin Scale (mRS) score at one 468 year was 0.60 (p<0.001), excluding mRS 6 (dead), and 0.88 (p<0.001) including mRS 6. We excluded 26 people without one-year mRS. There is a clear difference in median DAOH for mRS 0-2, and mRS 3-6. The edge of the boxes correspond to the first and third quartiles. The whiskers extend to the largest value no further than 1.5 * IQR from the edge. Individual outliers beyond the whiskers are not plotted due to data privacy reasons.

## Discussion

Our prospective, inception cohort of first-ever ICH in the Lothian region of Scotland demonstrates the persistent burden of ICH, with a rate of 28.1 (95%CI 27.6 – 28.5) per 100,000 per year adjusted to the UK 2024 population, a 41.2% (38.1% – 44.2%) 30-day case fatality, and 73.9% (68.8%–79.4%) dead or dependent at one year. HSD found similar incidence, survival and functional outcome after ICH, suggesting HSD could enable real-time monitoring of ICH burden, audit of ICH care, and research into ICH treatments.

Our incidence rate was similar to that in a recent systematic review of 29.2 per 100,000 person-years (95% CI 23.3-36.4). We furthermore found no time trend in incidence in a single, stable population.^2^ The Lothian population is aging,^17^ which may lead to increases in ICH incidence, but other factors could plausibly reduce incidence. Recent Scottish primary care initiatives to tackle hypertension (the strongest risk factor for ICH^18^) include the Quality Outcomes Framework, whereby from 2004 to 2015 Scottish primary care physicians received payments for monitoring and treating hypertension. Since 2015, people with hypertension in Scotland can send home blood pressure readings directly to their primary care physician through an app or SMS-messaging service.^19^ Other public health measures may reduce stroke incidence: reduction of dietary salt reduced blood pressure in England;^20^ the impact of minimum unit alcohol pricing in Scotland on cardiovascular disease is unexplored.

We had unique access to a reference standard of a prospective, population-based inception cohort to compare with HSD. Our results are consistent with hospital-based cohorts using ICD-9 codes in Italy, Norway and Finland, where the PPV of ICH coding was between 75% and 87%.^21^ Our PPV of 78.3% for ICH in HSD is lower than for ischemic stroke in similar studies; a study in UK Biobank found a PPV of 83% (74% – 90%) for ischemic stroke.^22^ However, the similar ICH incidence in HSD and LATCH suggests healthcare systems data are sufficiently accurate for monitoring incidence of ICH, without the expense or time-lag of a population-based cohort study. Our results support recent calls for use of routine data to monitor disease incidence.^23^ Overall, HSD detected more underlying medical conditions than LATCH, suggesting our methods of identification were more sensitive than criteria used by LATCH investigators.

Our study supports use of HSD for monitoring survival, with the caveat that disease only identified on death records should not be used to calculate survival. DAOH has previously been shown to correlate with age, comorbidities and stroke severity. ^24,25^ We have shown that, for people with ICH, DAOH correlates with a functional outcome measure, mRS, and may be a useful proxy for health service planning and clinical trials. HSD cannot currently reliably measure the full range of outcomes relevant to stroke survivors; examples include care use, depression or loss of income.

HSD has poor sensitivity for non-microvascular ICH. Prospective cohort studies allow detailed phenotyping, imaging, and pathological sample collection which is not yet possible in healthcare systems data. Pathological subtyping in HSD might be possible in future with AI analysis of brain imaging, imaging reports, or free text notes.^26^

Our study has several strengths. The prospective, hot-pursuit methods combined with retrospective methods employed in LATCH conform to standards recommended for stroke incidence studies.^27^ We undertook careful pathological subtyping to distinguish microvascular and other pathologies, which have different prognosis. The extended time period of the study contributes valuable data for trends in the burden of ICH both in the United Kingdom and worldwide.^2^ The HSD resource, DataLoch, includes primary care, secondary care, prescription and laboratory data, allowing extensive phenotyping.

## Limitations

Case identification for LATCH involved discussion with clinical teams. This may have influenced the hospital discharge letters and death certificates from which national hospital and death data are generated. LATCH participants did not routinely undergo angiographic imaging, which may mean some underlying macrovascular lesions were never identified.

The HSD resource, DataLoch, has 88% population coverage as not all primary care practices participate; we adjusted for this in our analysis. Nonetheless, the NHS provides universal healthcare and almost the entire Lothian population is registered with a primary care physician, reducing risk of ascertainment bias. Local demographic and socioeconomic factors may affect ICH incidence and outcome, and healthcare systems record data in varying ways worldwide; these factors affect the external validity of our results.

## Conclusions

We found a continuing burden of ICH over the time-period of the study. For ICH, we have provided a roadmap for using HSD for monitoring disease incidence and outcome, for both audit and research. ICH is an example of a rare disease which is only coded moderately well in HSD by non-medical coders; improvements could be achieved by using large language models to analyse and classify free-text notes.

## Data Sharing

The study used linked, de-identified, and aggregated individual patient level data from routine electronic healthcare data sources that is held in a trusted research environment by DataLoch (https://dataloch.org/). The study data and analysis code can be accessed by individuals who have undertaken the necessary governance training on application to DataLoch. The LATCH dataset is available from the authors upon reasonable request.

## Acknowledgement

Conflict of interest disclosures: AH is funded by a Medical Research Council (UK)/The Stroke Association fellowship (MR/Z504051/1). NS report grants from the Stroke Association (SA SCLM 23\100002) and Medical Research Council (G0900428). TM reports a grant from Scottish Heart and Arterial Risk Prevention (SHARP). JJML reports funding from the Wellcome Trust (214145/Z/18/Z), the UK Dementia Research Institute, British Heart Foundation (RE/24/130012), the Chief Scientist Office (PCL/25/12) and the Academy of Medical Sciences (SGCL033\1021). RASS reports grants from the British Heart Foundation and Chief Scientist Office Health Improvement, Medical Research Council, and Stroke Association. RASS is clinical director of the UK Registered Clinical Trials Unit Network. CS reports grants from the Medical Research Council and MAR from the Wellcome Trust. WNW is supported by the Chief Scientist Office (Scotland) and Health Data Research UK. All other authors have no conflicts of interest to declare. The funders had no role in the design, data collection, data analysis, and reporting of this study.

## Author contributions

AH, NS, WNW, and RASS contributed to the conception and design of the study. AH, NS, JJML, TJM, MAR, and RASS contributed to the acquisition of data. AH, WNW and RASS contributed to data analysis. All authors reviewed and edited the manuscript and approved the final version of the manuscript.

Dr Alice Hosking had full access to all the data in the study and takes responsibility for the integrity of the data and the accuracy of the data analysis.

